# Changes in SARS-CoV-2 Antibody Responses Impact the Estimates of Infections in Population-Based Seroprevalence Studies

**DOI:** 10.1101/2020.07.14.20153536

**Authors:** Craig Fenwick, Antony Croxatto, Alix T. Coste, Florence Pojer, Cyril André, Céline Pellaton, Alex Farina, Jérémy Campos, David Hacker, Kelvin Lau, Berend-Jan Bosch, Semira Gonseth Nussle, Murielle Bochud, Valerie D’Acremont, Didier Trono, Gilbert Greub, Giuseppe Pantaleo

## Abstract

We have determined SARS-CoV-2-specific antibody responses in a cohort of 96 individuals with acute infection and in 578 individuals enrolled in a seroprevalence population study in Switzerland including three groups, i.e. subjects with previous RT-PCR confirmed SARS-CoV-2 infections (n=90), ‘positive patient contacts’ (n=177) and ‘random selected subjects’ (n=311). SARS-CoV-2 antibody responses specific to the Spike (S), in the monomeric and native trimeric forms, and/or the nucleocapsid (N) proteins were equally sensitive in the acute infection phase. Interestingly, as compared to anti-S antibody responses, those against the N protein appear to wane in the post-infection and substantially underestimated the proportion of SARS-CoV-2 infections in the groups of ‘patient positive contacts’, i.e. 10.9 to 32.2% reduction and in the ‘random selected’ general population, i.e. up to 45% reduction. The overall reduction in seroprevalence targeting only anti-N IgG antibodies for the total cohort ranged from 9.4 to 31%. Of note, the use of the S protein in its native trimer form was more sensitive as compared to monomeric S proteins.

These results indicate that the assessment of anti-S IgG antibody responses against the native trimeric S protein should be implemented to estimate SARS-CoV-2 infections in population-based seroprevalence studies.

## Introduction

The SARS-CoV-2 is currently causing a devastating pandemic with more than 12.7 million documented infections and more than 566’000 deaths, according to the latest WHO situation report from July 13th, 2020.^1^ However, the true incidence of the infection is largely underestimated, since in most countries asymptomatic and paucisymptomatic people are tested only if they came in direct contact with sick patients or belong to at-risk subgroups. Therefore, it is a public health urgency to perform large-scale population-based studies in order to determine rates of seroprevalence during the first wave of the SARS-CoV-2 pandemic and to implement continued surveillance with the combined use of viral detection tests such as RT-PCR and serological testing. Seroprevalence studies are also instrumental to determine the proportion of individuals with potential protective immunity.^2-4^

SARS-CoV-2 antibody responses are characterized through the detection of IgG, IgA, and/or IgM. Detecting both IgA and IgG may increase sensitivity, particularly for people experiencing paucisymptomatic or asymptomatic infection. However, IgM does not seem to be of great benefit to overall sensitivity, since IgM appearance coincides with IgG antibodies during the early phase of infection, i.e. less than 15 days after the onset of symptoms and may increase the likelihood of false positive results due to cross-reactivity.^5,6^

SARS-CoV-2-specific antibody responses target two proteins: the nucleocapsid protein (N) and the Spike protein (S). It has been suggested that IgG antibodies targeting the S protein are more specific while those targeting at N may be more sensitive, particularly in the early phase of infection.^7^

However, the increased sensitivity of anti-N antibody response might be at the expense of specificity, given the relatively high protein sequence similarity of the N protein of SARS-CoV-2 with nucleocapsid proteins of other Coronaviridae and other viruses. Moreover, during the SARS outbreak (2002-2004), Chia et al observed that anti-N antibodies waned earlier than anti-S antibodies.^8^ Thus, anti-S antibody response might be more specific and circumvent a possible decrease of antibodies, previously observed with N protein of the SARS virus. Furthermore, it is still unknown the durability of SARS-CoV-2 antibody response. Previous studies have shown early disappearance of antibodies to SARS-associated coronavirus after recovery^9^ while other studies have shown longer durability of the antibody response.^10-13^

In the present study we have investigated SARS-CoV-2 antibody responses, both IgA and IgG in a cohort of 96 patients with moderate to severe symptoms during the first 33 days of the acute phase of infection and in a cohort of 578 subjects mostly paucisymptomatic and/or asymptomatic enrolled in a population-based seroprevalence study of the Vaud Canton in Switzerland. Antibody responses targeting either the N and/or the S proteins were investigated. Anti-S antibody responses were determined against monomeric moieties of the S1 protein and/or the native S trimeric form. Antibody responses against the S and N proteins were equally sensitive during the acute phase of infection while anti-N antibody responses waned in the post-infection phase. Importantly, the use of the trimeric as compared to the monomeric form of the S protein is associated with greater sensitivity in the detection of SARS-CoV-2 IgG antibody response in both the acute and post-infection phases.

Taken together, these results indicate that antibody responses against the native trimeric S protein should be used as a reference in population-based seroprevalence studies to provide more accurate estimates of SARS-CoV-2 infections in the general population.

## Results

### Antibody responses against the native trimeric versus the monomeric S protein

A stabilized trimer of the full-length S protein, encompassing both its S1 and S2 moieties, was coupled to beads for capturing antibodies in a new Luminex assay. We hypothesized that conformational epitopes would be preserved in the trimeric S protein, providing a greater sensitivity to detect IgG antibodies (**Supplementary Figure 1A and B**).^14^ First, the specificity for IgG antibody binding was established with sera from 256 pre-COVID-19 pandemic healthy adults from 18 to 81 years of age and an additional set of 108 patients (**Figure 1A**), which included: pregnant women, individuals infected with alphacoronaviruses (NL63 and 229E), betacoronaviruses (OC43 and HKU1), HIV, Rubella, HSV1, HSV2, RSV, CMV, EBV, influenza or varicella, as well as patients suffering from autoimmune diseases such as Lupus. The signal distribution for all SARS-CoV-2 negative sera was similar for the 256 pre-COVID-19 healthy adults and for the diverse panel of 108 subjects. A cut-off for positivity was set at 4-fold above a negative control standard, which is slightly more than four standard deviation above the mean of all negative control samples (mean MFI ratio 0.84 + 4×0.75 SD). Using this threshold, only one sera of the 256 pre-COVID-19 people and two patients with acute HIV or CMV viral infections gave a positive signal (**Figure 1 A**). As such, the Luminex assay using the stable trimeric S protein gave a high overall specificity of 99.2% and no cross-reactive antibodies were detected in sera from people infected with pre-pandemic coronaviruses or from patients with autoimmune diseases that can produce polyreactive antibodies.

**Figure 1:**
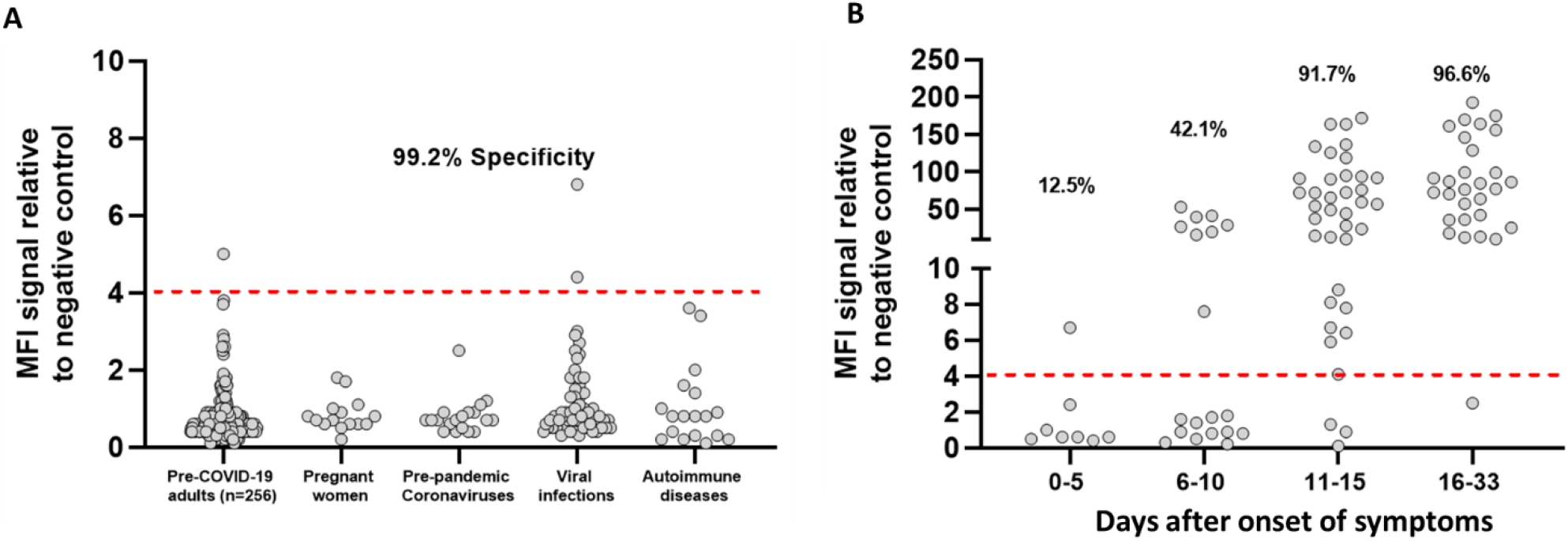
SARS-CoV-2-specific IgG binding antibody responses against the native trimeric S protein in a Luminex binding assay. Luminex beads covalently coupled with SARS-CoV-2 S protein trimer were used to monitor IgG binding antibody responses in pre-COVID-19 pandemic negative control sera and sera from SARS-CoV-2 PCR positive donors. MFI signals for serum antibody binding was expressed as a ratio compared to a negative control pool of pre-COVID-19 pandemic human serum tested in parallel. A) Assay specificity was evaluated using the sera from pre-COVID-19 pandemic healthy adults (n=256; ages ranging between 18 to 81 years of age), pregnant woman, pre-pandemic coronavirus infected donors (OC43, E229, NL63 and HKU1), patients with infectious diseases (HIV, Rubella, HSV1, CMV, EBV, influenza and varicella) and patients with autoimmune diseases including Lupus. B) The sensitivity of the S protein trimer was evaluated with sera from acute infected SARS-CoV-2 PCR-positive donors at 0-5 days, 6-10 days, 11-15 days and 16-33 days post-onset of symptoms. The red dashed line in A and B corresponds to the 4.0 cut-off for positivity in the IgG Luminex assay that was established by using mean value + 4×SD of all 364 pre-COVID-19 pandemic serum samples shown in A.

The sensitivity of the assay was next evaluated using sera from 96 acutely infected SARS-CoV-2 PCR-positive patients with blood sampling at 0-5 days, 6-10 days, 11-15 days and 16-33 days post-onset of symptoms (POS). As anticipated, sera collected during the early stage of the infection (0-5 days POS) had low or undetectable levels of anti-S protein IgG antibodies, with a rate of positivity of 12.5% (1 in 8 subjects; **Figure 1B**). Seropositivity increased to 42.1% (8/19) at 6-10 days POS and to 91.7% (33/36) at 11-15 days POS. Almost all patients with symptoms for 16-33 days (28/29; 96.6%) displayed high antibody titers for the S protein trimer. Interestingly, the only subject that was negative in the S protein trimer assay at day 25 post-onset of symptoms became seropositive when re-tested seven days later.

We then performed head-to-head comparisons of S trimeric versus S1 or RBD monomeric proteins for IgG antibody responses within the Luminex assay. The responses observed with the S monomeric proteins were similar in sensitivity to those described in previous studies ^15,16^ using monomeric proteins but inferior to those obtained with the trimeric S protein (**Supplemental Figure 2A and B**).

### Anti- IgA antibody response against the S protein trimer

We next evaluated the S protein trimer for the detection of anti-SARS-CoV-2 IgA antibodies. We established assay specificity and a cut-off threshold for positivity by screening sera from pre-COVID-19 healthy adults. Using four standard deviations above the standard negative control, this assay provided a 98.5% specificity in the 256 sera tested. The sensitivity was estimated on 81 out of 96 acute infected SARS-CoV-2 patients’ sera with positive detections ranging from 33.3% of patients at 0-5 days POS with seropositivity increasing to 68.8% in patients from the 6-10 days group. At 11-15 and 16-33 days POS, IgAs were detected in 94.4% and 90% of the cases, respectively (**Figure 2**).

**Figure 2:**
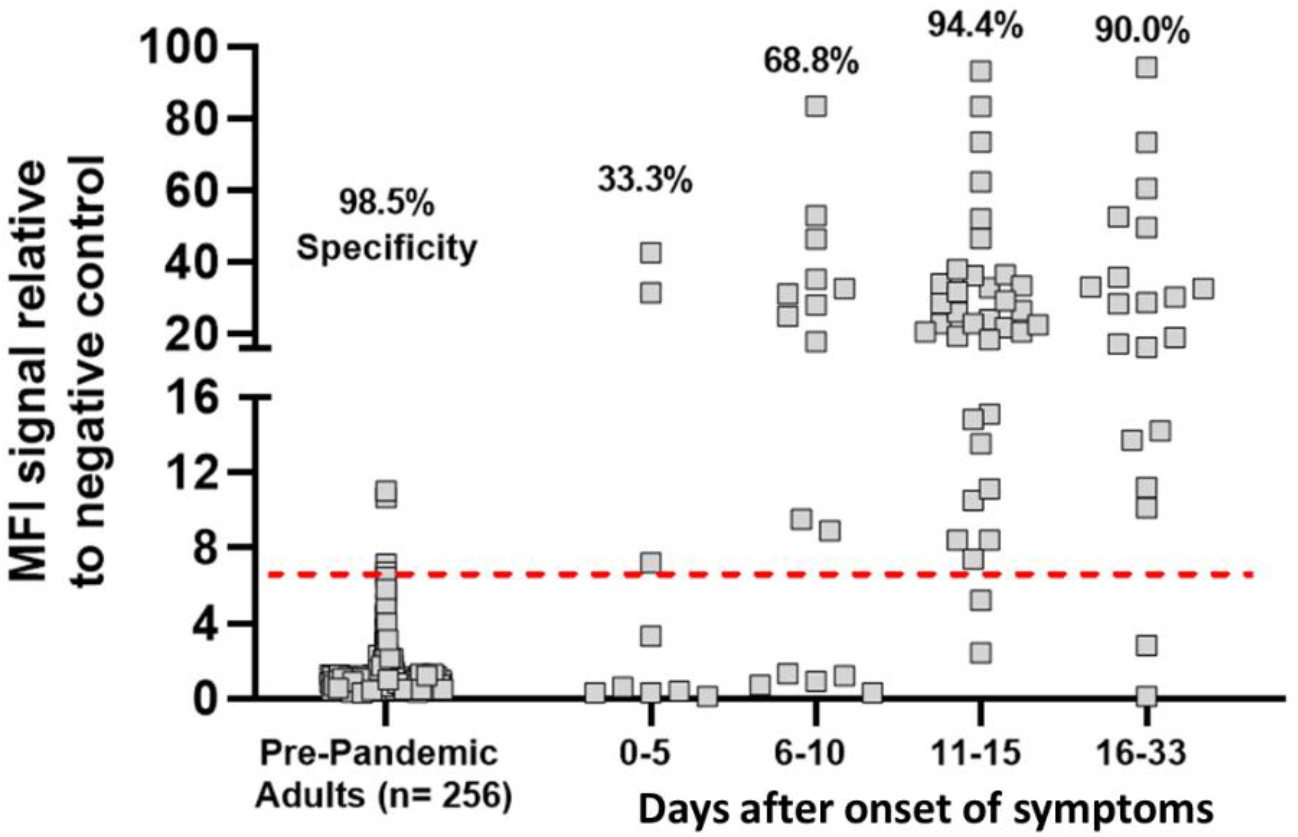
SARS-CoV-2-specific IgA binding antibody responses against the native trimeric S protein in a Luminex binding assay. The native trimeric S protein was used to monitor IgA binding antibodies in sera from pre-COVID-19 pandemic negative control donors and sera from acute SARS-CoV-2 PCR positive donors. The Luminex assay exhibited high specificity of 98.5% against a cohort of negative control donors and was effective at detecting IgA antibodies specific for S protein in most subjects in both the early stage (0 to 10 days) and later stage (11 to 33 days) after onset of symptoms in acute PCR-positive patients. The red dashed line corresponds to 6.5-fold MFI signal over the internal negative control and was established by using the mean value + 4×SD of the 256 pre-COVID-19 pandemic adult serum samples.

### Anti-S versus anti-N antibody responses during the acute phase of infection

Anti-S and anti-N antibody responses were determined using four different technologies, i.e Luminex, ELISA, CLIA and ECLIA. The different assays used were detecting the N protein alone, the N plus a monomeric antigen of the S protein, the monomeric S protein alone and the native trimeric S protein. More details about the five commercial assays used are contained in the Methods. We performed the comparison on the same set of 96 sera from patients with acute infection and stratified based on time between symptoms onset and sera collection as shown in **Figures 1B**. Small differences in the number of sera tested across assays is due to insufficient volume of some samples. The specificity was evaluated on a common panel of 65 pre-COVID-19 pandemic sera sampled before November 2019.

Increased sensitivity in the detection of both anti-N and anti-S IgG antibody responses was observed consistently over time post-symptoms regardless of the test used (**Figure 3 A-B, Table 1**). The use of the native trimeric S protein was associated with the higher sensitivity, i.e. detection of anti-S IgG antibodies in 97% of individuals tested > Day 15 POS, as compared to the use monomeric S and/or N proteins with a sensitivity ranging between 83 to 93% (**Figure 3 B-C, Table 1**). The specificity was equal or above 97% (**Figure 3 C**) regardless of the test and antigen used and none displayed cross-reactivity with sera from patients positive for 229E, OC43, HKU1, NL63 coronaviruses (**Figure 1A**).

**Table 1.** Cumulative data of SARS-CoV-2-specific IgG antibody responses on sera collected during the acute infection from hospitalized patients with moderate to severe symptoms

| Test | Days post-symptoms | N° sera from patients RT-PCR pos |  | IgG** Se | IgG** Se 95% CI | N° sera pre-COVID* |  | IgG** Sp | IgG** Sp 95% CI |
| --- | --- | --- | --- | --- | --- | --- | --- | --- | --- |
|  |  | Total | Positive in serology |  |  | Total | Positive in serology |  |  |
| Test 1 S Trimer | All | 94 | 71 | 0.76 | 0.66 to 0.83 | 65 | 0 | 1 | 0.94 to 1.0 |
|  | 0-5 | 8 | 1 | 0.13 | 0.0064 to 0.47 |  |  |  |  |
|  | 6-10 | 19 | 8 | 0.42 | 0.23 to 0.64 |  |  |  |  |
|  | 11-15 | 36 | 33 | 0.92 | 0.78 to 0.97 |  |  |  |  |
|  | 16-33 | 29 | 28 | 0.97 | 0.83 to 1.0 |  |  |  |  |
| Test 2 S1 mono | All | 96 | 49 | 0.51 | 0.41 to 0.61 | 65 | 0 | 1 | 0.95 to 1.0 |
|  | 0-5 | 8 | 0 | 0 | 0.0 to 0.32 |  |  |  |  |
|  | 6-10 | 19 | 4 | 0.21 | 0.085 to 0.43 |  |  |  |  |
|  | 11-15 | 37 | 21 | 0.57 | 0.41 to 0.71 |  |  |  |  |
|  | 16-33 | 29 | 24 | 0.83 | 0.65 to 0.92 |  |  |  |  |
| Test 3 S1 mono | All | 91 | 58 | 0.64 | 0.53 to 0.73 | 65 | 2 | 0.97 | 0.90 to 0.99 |
|  | 0-5 | 8 | 1 | 0.13 | 0.0064 to 0.47 |  |  |  |  |
|  | 6-10 | 18 | 5 | 0.28 | 0.12 to 0.51 |  |  |  |  |
|  | 11-15 | 34 | 27 | 0.79 | 0.63 to 0.90 |  |  |  |  |
|  | 16-33 | 29 | 25 | 0.86 | 0.69 to 0.95 |  |  |  |  |
| Test 4 N | All | 93 | 68 | 0.73 | 0.63 to 0.81 | 65 | 1 | 0.98 | 0.92 to 1.0 |
|  | 0-5 | 7 | 1 | 0.14 | 0.0073 to 0.51 |  |  |  |  |
|  | 6-10 | 18 | 8 | 0.44 | 0.25 to 0.66 |  |  |  |  |
|  | 11-15 | 36 | 32 | 0.89 | 0.75 to 0.96 |  |  |  |  |
|  | 16-33 | 29 | 26 | 0.89 | 0.74 to 0.96 |  |  |  |  |
| Test 5 N | All | 96 | 60 | 0.63 | 0.53 to 0.72 | 65 | 0 | 1 | 0.95 to 1.0 |
|  | 0-5 | 8 | 0 | 0 | 0.0 to 0.32 |  |  |  |  |
|  | 6-10 | 19 | 6 | 0.32 | 0.15 to 0.54 |  |  |  |  |
|  | 11-15 | 37 | 27 | 0.73 | 0.57 to 0.85 |  |  |  |  |
|  | 16-33 | 29 | 26 | 0.9 | 0.74 to 0.96 |  |  |  |  |
| Test 6 N | All | 96 | 68 | 0.73 | 0.62 to 0.80 | 65 | 0 | 1 | 0.95 to 1.0 |
|  | 0-5 | 8 | 0 | 0 | 0.0 to 0.32 |  |  |  |  |
|  | 6-10 | 19 | 9 | 0.47 | 0.27 to 0.68 |  |  |  |  |
|  | 11-15 | 37 | 32 | 0.86 | 0.72 to 0.94 |  |  |  |  |
|  | 16-33 | 29 | 27 | 0.93 | 0.78 to 0.99 |  |  |  |  |

Se : sensitivity ; Sp : specificity ; \* serum sampled before 1st November 2019 considered as “negative”; \*\*IgG, except for the Test 6 N, which corresponds to a pan-Ig assay

**Figure 3:**
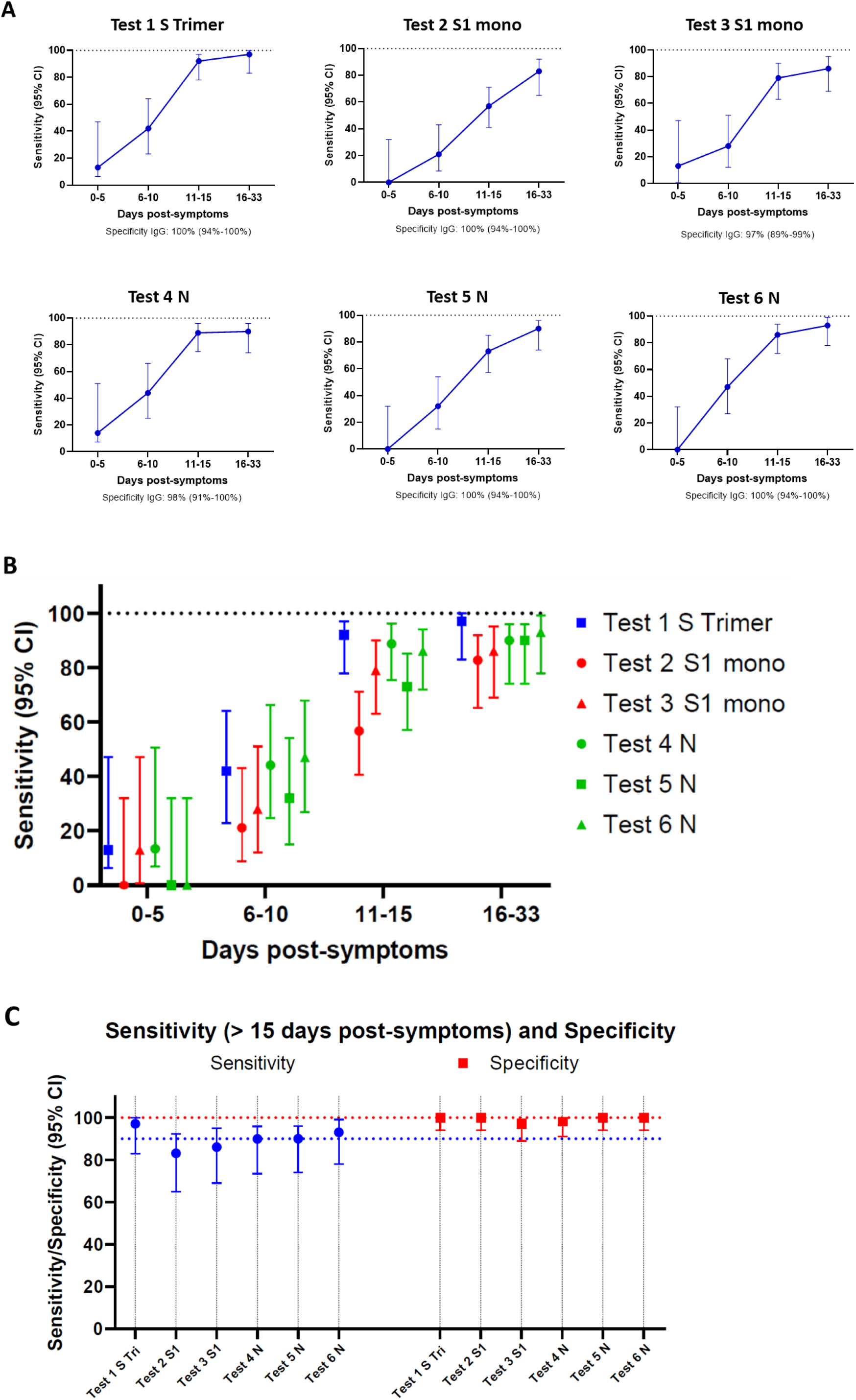
Comparative analysis of SARS-CoC-2-specific IgG binding antibody responses against S and N proteins in sera from patients with acute infection using six different serological tests. Sensitivity in detecting anti-SARS-CoV-2-specific IgG antibodies was assessed using the Luminex assay and five other commercial assay as described in the Methods. **A-B)** Serum samples were grouped by the number of days after initial onset of symptoms with sensitivity increasing over time. **C)** Comparison in sensitivity between the different tests in samples collected from day 16 to 33 post-symptoms. S Tri: trimeric S protein; S1: monomeric protein; N: nucleocapside protein.

Taken together, these results indicate that antibody responses targeting the S and/or the N proteins have similar sensitivity during the acute phase of infection.

### Comparison of anti-S and anti-N IgG antibody responses during the post-infection phase

We next evaluated anti-S and anti-N IgG antibody responses on 578 sera as part of a population-based seroprevalence study of the Vaud Canton in Switzerland, while being blind to the seropositivity status. These comparisons included 90 sera sampled from mildly to paucisymptomatic patients tested positive by RT-PCR, 177 sera sampled from ‘positive patient contacts’ of RT-PCR positive subjects, and 311 sera sampled from undefined, ‘random selected’ people from the general population aged 6 months and over. Results of the comparisons are shown in **Figure 4**, and **Table 2**. As expected, a good correlation in the proportion of seropositive individuals was observed between tests detecting antibody responses against the trimeric and/or monomeric S proteins while a poorer correlation was observed with those detecting anti-N antibody responses (**Figure 4**).

**Table 2:** Estimates of SARS-CoV-2 infections in the based population study.

|  | Test 1 S Trimer |  |  |  |  |  | Test 2 S1 mono |  |  |  |  |  | Test 3 S1 mono |  |  |  |  |  |
| --- | --- | --- | --- | --- | --- | --- | --- | --- | --- | --- | --- | --- | --- | --- | --- | --- | --- | --- |
|  | Positive RT-PCR |  | Positive patient contacts |  | Random selected Subjects |  | Positive RT-PCR |  | Positive patient contacts |  | Random selected Subjects |  | Positive RT-PCR |  | Positive patient contacts |  | Random selected Subjects |  |
|  | % | N | % | N | % | N | % | N | % | N | % | N | % | N | % | N | % | N |
| Positive | 96.7 | 87 | 36.2 | 64 | 6.4 | 20 | 88.9 | 80 | 28.2 | 50 | 4.5 | 14 | 88.9 | 80 | 29.9 | 53 | 4.2 | 13 |
| Negative | 2.2 | 2 | 62.7 | 111 | 90.7 | 282 | 8.9 | 8 | 70.6 | 125 | 95.2 | 296 | 6.7 | 6 | 68.9 | 122 | 94.9 | 295 |
| Limit | 1.1 | 1 | 1.1 | 2 | 2.9 | 9 | 2.2 | 2 | 1.1 | 2 | 0.3 | 1 | 4.4 | 4 | 1.1 | 2 | 1.0 | 3 |

|  | Test 4 N |  |  |  |  |  | Test 5 N |  |  |  |  |  | Test 6 N |  |  |  |  |  |
| --- | --- | --- | --- | --- | --- | --- | --- | --- | --- | --- | --- | --- | --- | --- | --- | --- | --- | --- |
|  | Positive RT-PCR |  | Positive patient contacts |  | Random selected Subjects |  | Positive RT-PCR |  | Positive patient contacts |  | Random selected Subjects |  | Positive RT-PCR |  | Positive patient contacts |  | Random selected Subjects |  |
|  | % | N | % | N | % | N | % | N | % | N | % | N | % | N | % | N | % | N |
| Positive | 76.7 | 69 | 26.0 | 46 | 4.5 | 14 | 71.1 | 64 | 24.3 | 43 | 3.5 | 11 | 95.6 | 86 | 32.2 | 57 | 3.9 | 12 |
| Negative | 17.8 | 16 | 70.6 | 125 | 93.6 | 291 | 28.9 | 26 | 75.7 | 134 | 96.5 | 300 | 4.4 | 4 | 67.8 | 120 | 96.1 | 299 |
| Limit | 5.6 | 5 | 3.4 | 6 | 1.9 | 6 | 0.0 | 0 | 0.0 | 0 | 0.0 | 0 | 0.0 | 0 | 0.0 | 0 | 0.0 | 0 |

Se Test 1 Tri CI 95% : 0.92 to 0.996 on Positive RT-PCR sera Se Test 2 S CI 95% : 0.83 to 0.95 Se Test 3 S CI 95% : 0.86 to 0.97 Se Test 4 N CI 95% : 0.72 to 0.88 Se Test 5 N CI 95% : 0.61 to 0.79 Se Test 6 N CI 95% : 0.89 to 0.98

**Figure 4:**
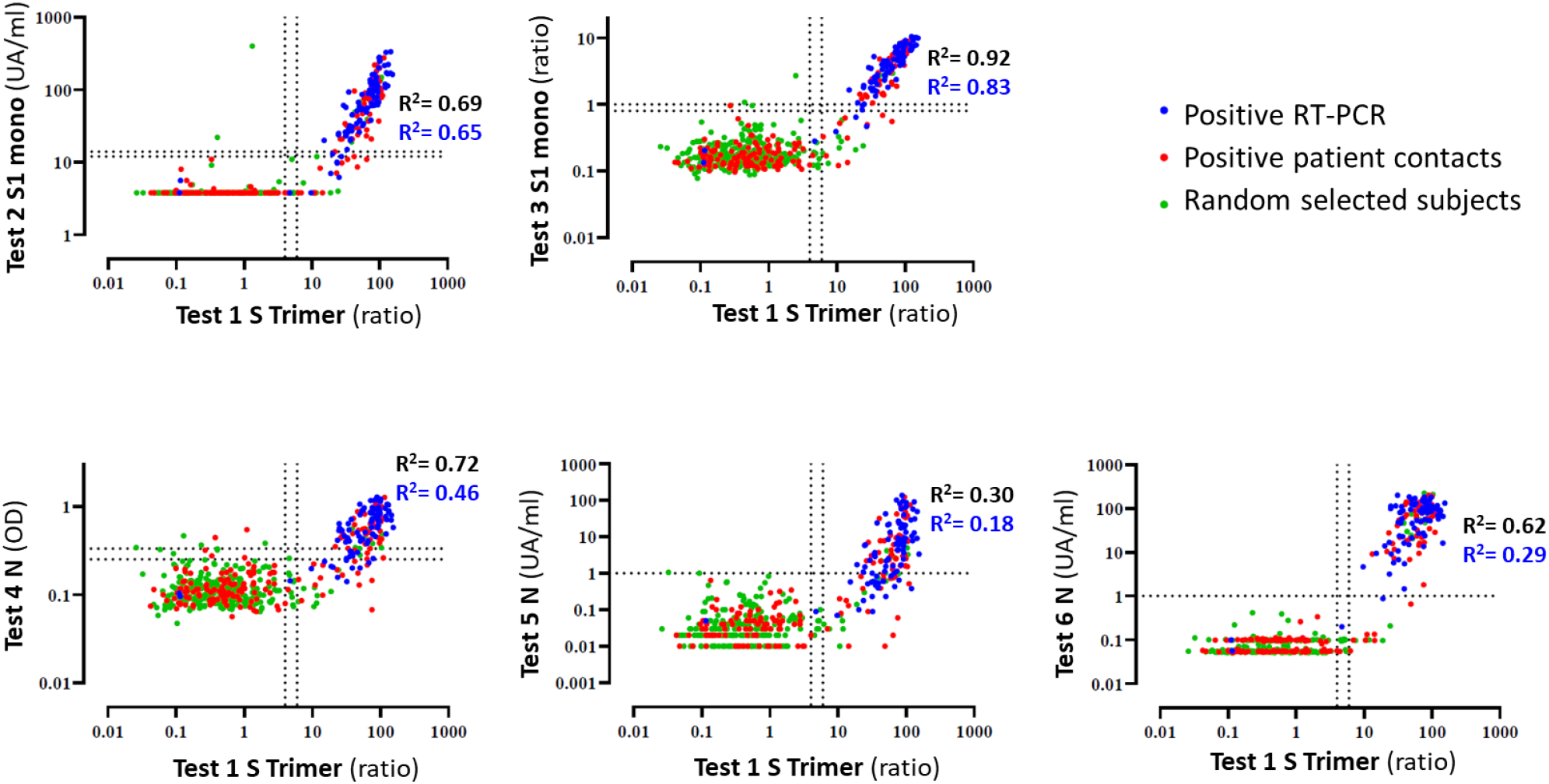
Comparative analysis of IgG antibody responses against the trimeric S protein versus monomeric S and/or N proteins. Signal intensities for the different subject sera in the post-infection cohort were compared between the Luminex and the five other serological assays. Collected sera were from patients with a documented positive SARS-CoV-2 RT-PCR (90 sera; blue dots), positive patient contacts with a SARS-CoV-2 RT-PCR positive patient (177 sera; red dots) and randomly selected individuals from the general population (311 sera; green dots). Pearson correlation R2 values are given for all 578 participants (black text) or for 183 Luminex positive sera (blue text).

With regard to the ‘RT-PCR positive’ group (n=90 individuals) (**Figure 4**, blue dots), the best sensitivity (96.7%) was found with the use of the trimeric S protein as compared to that of monomeric S and N proteins (**Table 2**).

Regarding ‘positive patient contact’ group (**Figure 4**, red dots), the highest positivity rate (36.2%) was observed with the trimeric S protein while positivity rates ranged between 32.2 and 24.3% with the other antigenic proteins (**Figure 4 and Table 2**).

With regard to the ‘random selected’ group, (**Figure 4**, green dots; **Table 2**), we observed that anti-S antibody responses identified greater percentages of SARS-CoV-2 positive people (between 6.4 to 4.2%), than anti-N antibody responses (4.5 to 3.5%). Importantly, the pan-Ig test (#6 in **Table 2**) using the N protein antigen was the second most sensitive assay in the acute infected cohort and in the ‘RT-PCR positive’ and ‘positive patient contacts’ groups, but conversely, was one of the least sensitive test (3.9%) in detecting seropositive people randomly selected from the general population. The significantly higher sensitivity of the trimeric S protein antigen in the post-infection setting is highlighted in **Figure 5**. Compared to the N and or monomeric S antigens, the trimeric S protein identified 10.9% to 32.8% more positive subjects in the ‘positive patient contacts’ group, 30% to 45% more positive subjects in the ‘random selected’ group and 17.9% to 35.7% more positive subjects in a combined analysis of the ‘positive patient contacts’ and ‘random selected’ groups. In the overall post-infection cohort of 578 subjects, the trimeric S protein performed significantly better and detected between 9.4% and 31% more seropositive participants than the N and/or the S monomeric proteins (**Figure 5**).

**Figure 5:**
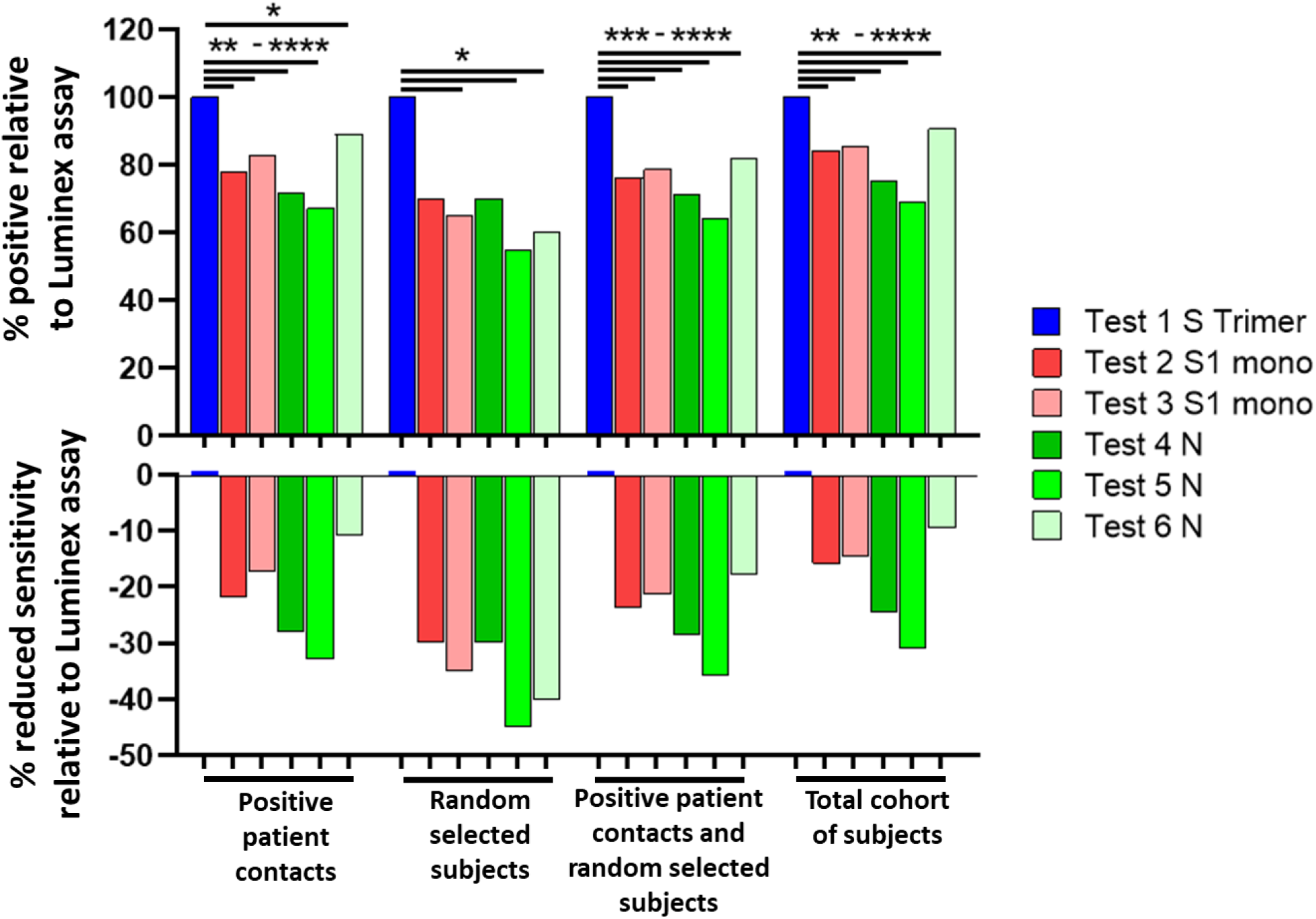
SARS-CoV-2-specific antibody responses to the trimeric S protein have significantly increased sensitivity as compared to S1 monomeric and/or N proteins in the post-infection population-based study. Analysis shows the percentage of seropositive subjects relative to the estimates obtained with the trimeric S protein (top) and the percentage of reduced sensitivity relative to the S1 monomeric and/or N proteins (bottom). Tests with blue bars used the S protein trimer as their bait for binding serum antibodies while the red bars used the monomeric S1 protein and the green bars the N protein. Statistical analysis was performed using the McNemar test for matched participant samples where P<0.045 (*); P<0.0022 (**); P<0.0009 (***); P≤0.0001 (****).

Taken together, these results indicate that anti-N antibody responses may substantially (i.e. 30% to 45%) underestimate the proportion of SARS-CoV-2 exposed individuals compared to anti-S antibody responses in population-based seroprevalence studies.

## Discussion

Population-based seroprevalence studies are important to monitor the dynamics of the pandemic, to have a better appreciation of the number of infections and to determine the proportion of the population that has developed specific SARS-CoV-2 immunity. Population-based seroprevalence studies performed in Switzerland, Spain and in New York City indicate that a minor percentage of the population, ranging from 10 to 20% of individuals, has been infected with SARS-CoV-2.^2-4,17-19^ The estimates of SARS-CoV-2 infected individuals from seroprevalence studies may be substantially influenced by qualitative and quantitative changes in the antibody responses from the transition from the acute to the post-infection phase, the clinical severity of the infection and the antigenic protein used for the detection of the antibody responses.

SARS-CoV-2 specific antibodies (predominantly IgG) targeting either the S or the N proteins are generally assessed in both the acute and post-infection phases. The majority of studies and the validation of the tests with regard to the sensitivity and specificity has been mostly performed on cohorts from patients during the acute phase of infection.^16,17,20,21^ The results from these studies indicate the use of N and S proteins were considered as equally sensitive, with a generally higher sensitivity for the N protein to monitor the development of antibody responses. Based on these observations in the acute phase of infection, it has been assumed that determination of antibody responses against the N or S proteins would be equally suitable in the post-infection phase for population-based seroprevalence studies. However, limited information is yet available on the evolution of the antibody response during the transition from the acute to the post-infection phase and, in particular, on the antibody responses against the two targets, S and N proteins. Furthermore, the population-based studies comprise diverse populations of individuals including RT-PCR positive individuals with moderate to severe symptomatic infection who required hospitalization, RT-PCR positive individuals with mild symptoms who did not require hospitalization and pauci-/asymptomatic individuals with no previous RT-PCR confirmation of COVID-19 infection. Previous studies have shown that the magnitude of the antibody response may be influenced by the severity of the symptoms with robust antibody response in patients with severe infection while weaker antibody response in patients with mild infection. Therefore, antibody responses can be lower in pauci-/asymptomatic individuals. For these reasons, population-based studies can be very challenging to estimate the proportion of SARS-CoV-2 infections in individuals who have experience pauci-/asymptomatic infection.

Our results indicate a substantial drop in the sensitivity of antibody responses specific to the N protein thus strongly suggesting a waning of these responses in the post-infection phase. In this regard, the estimated seroprevalence in the ‘positive patient contacts’ and ‘random selected’ groups is mostly impacted when only anti-N responses are assessed with an underestimation ranging from 11 to 33% for the former and 30 to 45% for the latter group as compared to anti-S trimeric responses within the same groups.

Of note, the underestimation of SARS-CoV-2 seropositive individuals was also observed for antibody responses against monomeric S1 or RBD and was in the range of 18-22% in the ‘positive patient contacts’ and 30-35% in the ‘random selected’ groups samples. The greater sensitivity of antibody responses found against the trimeric S protein likely results from antibodies binding to the S2 subunit and the conservation of conformational epitopes within the higher order structure. This increased sensitivity was not obtained at the expense of cross-reactivity, since the specificity observed using the trimeric S protein was >99%. Overall, the underestimation of SARS-CoV-2 seropositive individuals was less important in the ‘Positive RT-PCR patients’ group ranging from 1 to 26%.

A recent study^22^ has shown that 40% of asymptomatic individuals became seronegative over time. However, anti-N antibody responses were determined in this study. Based on our results, it is likely that the loss of antibody response observed is due to the selective waning of the anti-N rather antibody responses rather than to a global reduction of the SARS-CoV-2 antibody response.

Furthermore, the present findings are also important for the development appropriate monitoring strategies for the evaluation and development of vaccines against SARS-CoV-2.

In conclusion, these results provide new insights in the evolution of the SARS-CoV-2 antibody response from the acute to the post-infection phase and indicate that the detection of antibody responses against the native trimeric S protein should be implemented to avoid large underestimation of SARS-CoV-2 infections in population-based seroprevalence studies.

## Material and Methods

### Study populations

#### Patients with acute infections

Comparison of tests for acute/sub-acute phase of the infection was performed on 161 sera, including i) 96 sera, expected to be positive, sampled from hospitalized patients with severe to mild symptoms 0 to 45 days post onset of the symptoms and documented with a positive SARS-CoV-2 RT-PCR ; ii) 65 sera, expected to be negative, sampled before November 2020, presented as pre-COVID-19 sera, and including 18 samples from patient documented positive for a Human coronavirus (E229, OC43, HKU1, NL63) RT-PCR. Date of the symptoms onset were extracted from the electronic record of the 96 SARS-CoV-2 RT-PCR positive patients.

#### Post-infection cohort

A second comparison of tests was performed on sera from the seroprevalence study of the Vaud Canton in Switzerland (SerocoViD) performed by the Centre for Primary Care and Public Health, University of Lausanne (Unisanté). Out of the 1,942 participants who provided a blood sample between May 4 and June 27, 2020, a subset of 578 subjects were included in the present analysis, of which: i) 90 subjects were expected to be positive—sampled from mostly mildly to paucisymptomatic patients (only 21% had been hospitalized) documented with a positive SARS-CoV-2 RT-PCR ; ii) 177 were sampled from contacts of RT-PCR positive subjects, and iii) 311 were randomly selected subjects in the general population. There were 304 women (52.6%), and the mean age was 39.2 years (SD 24.2, range: 6 months to 90 years).

#### Pre-COVID-19 pandemic donors

Negative control serum samples from 256 adult healthy donors with ages ranging for 18 to 81 years of age were collected prior to November 2019 as part of the Swiss Immune Setpoint study sponsored by Swiss Vaccine Research Institute. Specificity tests for the Luminex S-protein assay with a diverse set of 108 patient sera included the 65 sera collected prior to November 2019 and used in the blinded tested performed with all six assays and an additional 43 patient samples. This diverse set of samples consisted of sera from pregnant women (n=14), pre-pandemic coronavirus infected donors (OC43, E229, NL63 and HKU1; n=19), patients with infectious diseases (HIV, Rubella, HSV1, HSV2, CMV, EBV, influenza and varicella; n=57) and patients with autoimmune diseases including Lupus (n=18). Study design and use of subject sera samples were approved by the Institutional Review Board of the Lausanne University Hospital and the ‘Commission d’éthique du Canton de Vaud’ (CER-VD) stated that authorization was not required.

#### Preparation of Luminex beads

Luminex beads used for the serological binding assays were prepared by covalent coupling of SARS-CoV-2 proteins with MagPlex beads using the manufacture’s protocol with a Bio-Plex Amine Coupling Kit (Bio-Rad, France). Briefly, 1 ml of MagPlex-C Microspheres (Luminex) were washed with wash buffer and then resuspended in activation buffer containing a freshly prepared solution of 1-ethyl-3-(3-dimethylaminopropyl) carbodiimide (EDC) and N-hydroxysulfosuccinimide (S-NHS), (ThermoFischer, USA). Activated beads were washed in PBS followed by the addition of 50 μg of protein antigen. The coupling reaction was performed at 4 °C overnight with bead agitation using a Hula-Mixer (ThermoFischer). Beads were then washed with PBS, resuspended in blocking buffer then incubated for 30 minutes with agitation at room temperature. Following a final PBS washing step, beads were resuspended in 1.5 ml of storage buffer and kept protected from light in an opaque tube at 4 °C. Each of the SARS-CoV-2 proteins was coupled with different colored MagPlex beads so that tests could be performed with a single protein bead per well or in a multiplexed Luminex serological binding assay.

#### SARS-CoV-2 proteins evaluated in Luminex assay

The S protein trimer was designed to mimic the native trimeric conformation of the protein in vivo and the expression vector was kindly provided by Prof. Jason McLellan, University of Texas, Austin; 25. It encoded the prefusion ectodomain of the SARS-CoV-2 Spike with a C-terminal T4 foldon fusion domain to stabilize the trimer complex along with C-terminal 8x His and 2x Strep tags for affinity purification. The trimeric Spike protein was transiently expressed in suspension-adapted ExpiCHO cells (Thermo Fisher) in ProCHO5 medium (Lonza) at 5 ×106 cells/mL using PEI MAX (Polysciences) for DNA delivery. At 1 h post-transfection, dimethyl sulfoxide (DMSO; AppliChem) was added to 2% (v/v). Following a 7-day incubation with agitation at 31 °C and 4.5% CO2, the cell culture medium was harvested and clarified using a 0.22 µm filter. The conditioned medium was loaded onto Streptactin (IBA) and StrepTrap HP (Cytiva) columns in tandem, washed with PBS, and eluted with 10 mM desthiobiotin in PBS. The purity of S protein trimer was determined to be > 99% pure by SDS-PAGE analysis.

Receptor binding domain (RBD) and S1 SARS-CoV-2 proteins were prepared as previously described.^23^ In initial characterization of the assays, serum dilutions of 1/50 down to 1/ 2’700 were evaluated for SARS-CoV-2 PCR-positive subjects and healthy donors. A 1/300 dilution of serum was selected for screening patient samples since it showed a high MFI signal for all donors and a low background staining with serum samples from pre-COVID-19 pandemic healthy donors. In addition to the high positive signal and low background, <1 µl of serum was needed to evaluate anti-SARS-CoV-2 antibody binding in the Luminex assay binding assays.

#### Luminex anti-SARS-CoV-2 antibody binding assay

Luminex beads coupled with the Spike, RBD or S1 proteins were diluted 1/100 in PBS with 50 µl added to each well of a Bio-Plex Pro 96-well Flat Bottom Plates (Bio-Rad). Following bead washing with PBS on a magnetic plate washer (MAG2x program), 50 µl of individual serum samples diluted at 1/300 in PBS, were added to the plate wells. Along with samples, three replicates of a 1/300 negative control pool of pre-COVID-19 pandemic healthy human sera (BioWest human serum AB males; VWR) were evaluated on each 96-well plate. Plates were sealed with adhesive film, protected from light with a dark cover and agitated at 500 rpm for 60 minutes on a plate shaker. Beads were then washed on the magnetic plate washer and anti-human IgG-PE secondary antibody (OneLambda ThermoFisher) was added at a 1/100 dilution with 50µl per well. Plates were agitated for 45 minutes, and then washed on the magnetic plate washer. Beads resuspended in 80 µl of reading buffer were agitated 5 minutes at 700 rpm on the plate shaker then read directly on a Luminex FLEXMAP 3D plate reader (ThermoFisher). MFI signal for each test serum samples was divided by the mean signal for the negative control samples to yield an MFI ratio that normalized values between plates and between different Luminex instruments tested. Considering that two of the three false positives from the 364 SARS-CoV-2 negative donors had MFI signals less than 6 (**Figure 1A**), an additional criteria for positivity was established for large general population screens, including the post-infection cohort. Here, sera with signal intensities between 4 and 6 were defined as being at the limit of positivity, which increases the assay sensitivity to 99.7% with only one acute HIV infected subject having a 6.8 MFI signal.

#### Immunoassays

The new Luminex S protein trimer IgG assay was compared with five commercially available SARS-CoV-2 immunoassays: i) two ELISAs from EuroImmun (Test 3 S1 mono) and Epitope Diagnostics (Test 4 N protein) detecting IgG against the S1 and N proteins, respectively, ii) two CLIA from Diasorin (Test 1 S1 mono) and Snibe (Test 5 N protein) detecting IgG against S1 protein and N protein + S antigen peptide, respectively, and iii) a pan-Ig ECLIA from Roche (Test 6 N protein) targeting the N protein. The Snibe assay was grouped with the N protein assays in our analysis since it contained only a portion of the S1 protein.

ELISA and CLIA were performed according to the manufacturers’ instructions. EuroImmun and Epitope Diagnostic IgG ELISA were done manually as per protocol with the exception of washing steps performed with a microplate washer (PW40, Bio-Rad, France). Optical densities (OD) was measured with a microplate reader (800 TSI, BioTek, USA). Each sample was measured in duplicates. The LIAISON® SARS-CoV-2 IgG kit was performed on a Liaison® XL (Diasorin, Italy), and the MAGLUMI(tm) 2019-nCoV IgG and IgM kits were performed on a MAGLUMI(tm) 800 (Snibe, China). The Elecsys anti-SARS-CoV-2 was performed on a COBAS 6000 (Roche, Switzerland).

#### Statistical analyses

The sensitivity of the different tests was calculated according to day post-symptoms on expected positive sera taken from patients with a positive RT-PCR. The RT-PCR was previously performed according to Corman et al.^24^ on our automated molecular diagnostic platform.^25^

Sensitivity and specificity of the tests with 95% CI (Wilson/Brown method of GraphPad Prism 8.3.0) were calculated with Excel and GraphPad prism. For comparisons between the Luminex assay and the five other serological assays, R^2^ values were calculated using the Pearson test and the McNemar’s test was used to determine the P-value significant differences for sensitivities in detecting seropositive subjects in the patient subsets within the post-infection cohort. All statistics were done with GraphPad prism.

## Data Availability

Data available upon request

## Supplemental Figures

**Supplementary Figure 1:**
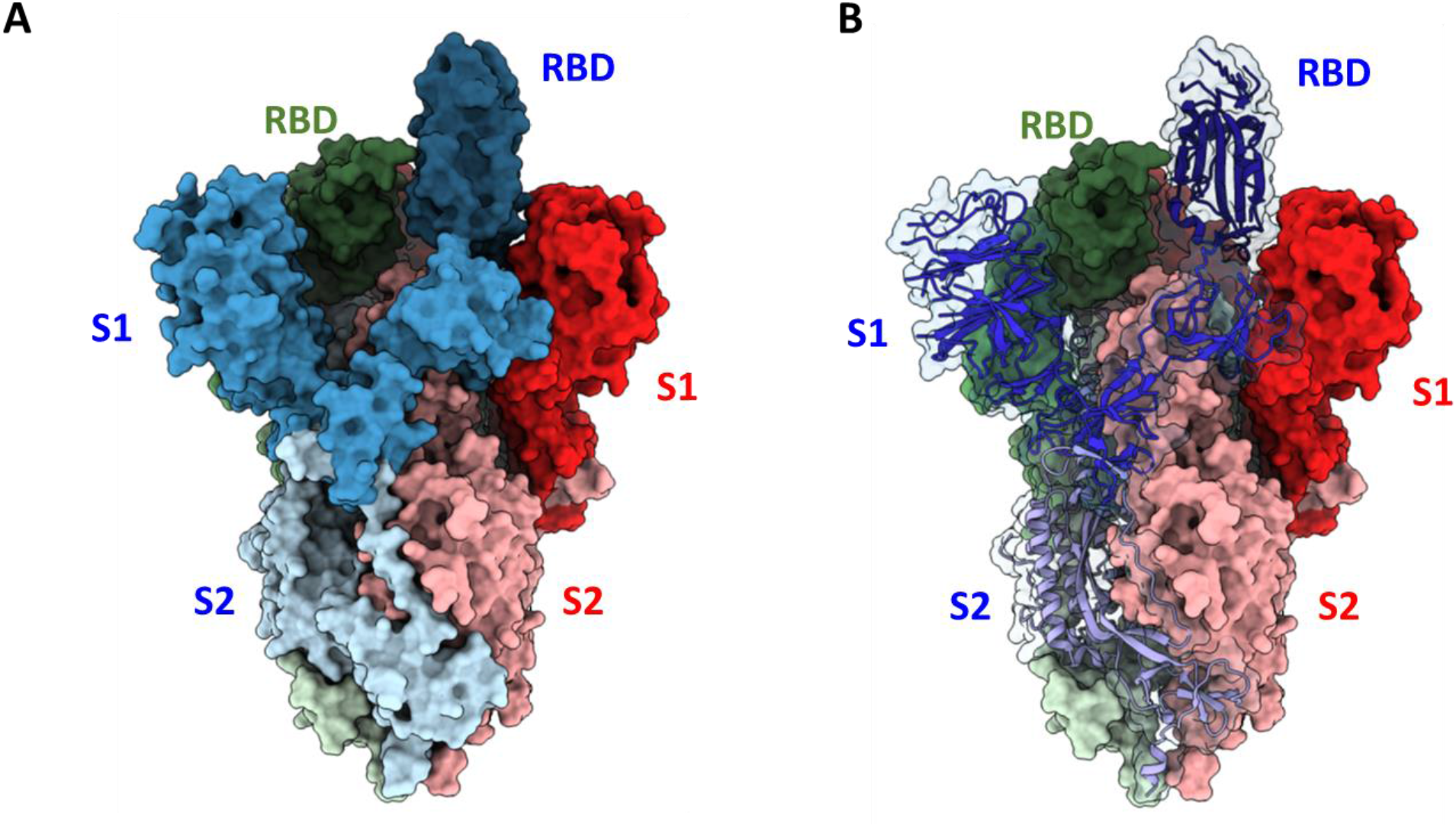
Structural representation of the SARS-CoV-2 S protein Trimer. A) Space filled representation of the S protein with trimer subunits shown in blue, red and green (PDB 6VSB). The labeled S2, S1 and RBD portions of each subunit are in light, mid and dark colors, respectively. Compared to the monomeric S1 protein, this image demonstrates that the native S protein trimer, consisting of S1 and S1 proteins, that has significantly greater conformational epitopes for antibody binding that are only present in the higher order structure. B) S protein trimer with the blue subunit represented as a ribbon structural.

**Supplementary Figure 2:**
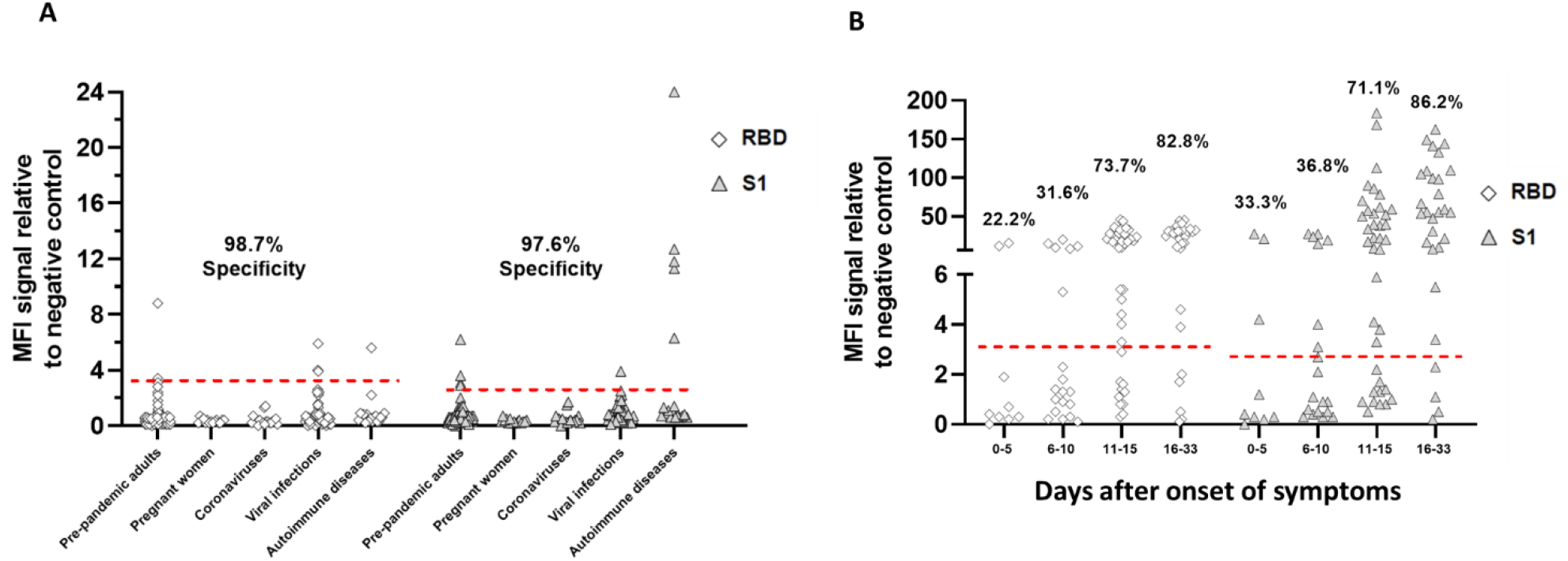
SARS-CoV-2-specific IgG binding antibody responses against the monomeric S1 and RBD domains in a Luminex binding assay. A) Specificity was assessed using the cohort of negative control sera described in Figure 1A. B) Sensitivity was determined with the 94 acute infected serum samples described in Figure 1B. The cut-off for positivity used in the RBD and S1 Luminex IgG assays are 3.2- and 2.8-fold over the negative control, respectively and were established by using mean value + 4×SD of each for the 364 pre-COVID-19 pandemic serum samples in A.

